# Population immunity of natural infection, primary-series vaccination, and booster vaccination in Qatar during the COVID-19 pandemic: An observational study

**DOI:** 10.1101/2023.04.28.23289254

**Authors:** Suelen H. Qassim, Hiam Chemaitelly, Houssein H. Ayoub, Peter Coyle, Patrick Tang, Hadi M. Yassine, Asmaa A. Al Thani, Hebah A. Al-Khatib, Mohammad R. Hasan, Zaina Al-Kanaani, Einas Al-Kuwari, Andrew Jeremijenko, Anvar Hassan Kaleeckal, Ali Nizar Latif, Riyazuddin Mohammad Shaik, Hanan F. Abdul-Rahim, Gheyath K. Nasrallah, Mohamed Ghaith Al-Kuwari, Adeel A. Butt, Hamad Eid Al-Romaihi, Mohamed H. Al-Thani, Abdullatif Al-Khal, Roberto Bertollini, Laith J. Abu-Raddad

**Author notes:** Correspondence: Dr. Suelen H. Qassim and Professor Laith J. Abu-Raddad, Infectious Disease Epidemiology Group, World Health Organization Collaborating Centre for Disease Epidemiology Analytics on HIV/AIDS, Sexually Transmitted Infections, and Viral Hepatitis, Weill Cornell Medicine - Qatar, Qatar Foundation - Education City, P.O. Box 24144, Doha, Qatar.

## Abstract

**Background:** Waning of natural infection protection and vaccine protection highlight the need to evaluate changes in population immunity over time. Population immunity of previous SARS-CoV-2 infection or of COVID-19 vaccination are defined, respectively, as the overall protection against reinfection or against breakthrough infection at a given point in time in a given population.

**Methods:** We estimated these population immunities in Qatar’s population between July 1, 2020 and November 30, 2022, to discern generic features of the epidemiology of SARS-CoV-2. Effectiveness of previous infection, mRNA primary-series vaccination, and mRNA booster (third-dose) vaccination in preventing infection were estimated, month by month, using matched, test-negative, case-control studies.

**Findings:** Previous-infection effectiveness against reinfection was strong before emergence of Omicron, but declined with time after a wave and rebounded after a new wave. Effectiveness dropped immediately after Omicron emergence from 88.3% (95% CI: 84.8-91.0%) in November 2021 to 51.0% (95% CI: 48.3-53.6%) in December 2021. Primary-series effectiveness against infection was 84.0% (95% CI: 83.0-85.0%) in April 2021, soon after introduction of vaccination, before waning gradually to 52.7% (95% CI: 46.5-58.2%) by November of 2021. Effectiveness declined linearly by ∼1 percentage point every 5 days. After Omicron emergence, effectiveness dropped suddenly from 52.7% (95% CI: 46.5-58.2%) in November 2021 to negligible levels in December 2021. Booster effectiveness dropped immediately after Omicron emergence from 83.0% (95% CI: 65.6 -91.6%) in November 2021 to 32.9% (95% CI: 26.7-38.5%) in December 2021, and continued to decline thereafter. Effectiveness of previous infection and vaccination against severe, critical, or fatal COVID-19 were generally >80% throughout the study duration.

**Interpretation:** High population immunity may not be sustained beyond a year. This creates fertile grounds for repeated waves of infection to occur, but these waves may increasingly exhibit a benign pattern of infection.

**Funding:** The Biomedical Research Program and the Biostatistics, Epidemiology, and the Biomathematics Research Core, both at Weill Cornell Medicine-Qatar, Ministry of Public Health, Hamad Medical Corporation, Sidra Medicine, Qatar Genome Programme, Qatar University Biomedical Research Center, and Qatar University Internal Grant ID QUCG-CAS-23/24-114.

**Research in context:** *Evidence before this study:* SARS-CoV-2 infection induces protection against reinfection, but this protection wanes with time since last infection. Similarly, COVID-19 primary-series and booster vaccination induce protection against SARS-CoV-2 infection, but this protection also wanes with time since last dose. These immunity patterns demonstrate the need for the concept of *population immunity* to track evolution of overall immune protection over time in a given population. Previous-infection and vaccine population immunities in a specific country can be defined as the overall protection against infection at a given point in time in the full national population. A search of PubMed, Google Scholar, and the International Vaccine Access Center’s VIEW-hub databases up to April 21, 2023 using the keywords “vaccination”, “infection”, “immunity”, “protection”, “SARS-CoV-2”, and “COVID-19” did not identify studies that investigated this epidemiological concept for a national population throughout the COVID-19 pandemic.

*Added value of this study:* This study analyzed the national federated databases for SARS-CoV-2 infection and COVID-19 vaccination in Qatar, a country that experienced SARS-CoV-2 waves dominated by different pre-Omicron variants and Omicron subvariants. Using a matched, test-negative study design, population immunity against infection of each of previous infection, primary-series vaccination, and booster vaccination were characterized at the national level month by month for two calendar years to discern generic features of the epidemiology of SARS-CoV-2. The three forms of population immunity showed rapid variation over time driven by waning of protection. Vaccine-derived population immunity declined by 1 absolute percentage point every 5 days. Omicron introduction immensely reduced the three forms of population immunity within one month by about 50 absolute percentage points. Meanwhile, previous-infection and vaccine population immunities against severe COVID-19 were durable with slow waning even after Omicron emergence.

*Implications of all available evidence:* Both previous-infection and vaccine population immunities vary rapidly at a national level creating fertile grounds for repeated waves of infection to occur even within months of each other. High levels of population immunity may not be sustained for more than a year or so. Preventing infection/reinfection, transmission, or future waves of infection cannot sustainably be done with current vaccines nor by the entire population being infected. Timely administration of boosters for those vulnerable to severe COVID-19 may remain essential for years to come. Repeated waves of infection may also facilitate further evolution of the virus and continual immune evasion. Emergence of a new variant that is substantially different from circulating variants can suddenly and immensely reduce population immunity leading to large epidemic waves. However, the durability of population immunity against severe COVID-19 will likely curtail the severity of future waves.

## Introduction

Although immune protection of the primary series of coronavirus disease 2019 (COVID-19) vaccines is high against severe acute respiratory syndrome coronavirus 2 (SARS-CoV-2) infection immediately after the second dose,^1, 2^ protection wanes with time and may not last for more than 1 year after the second dose.^3–5^ The emergence of the immune-evasive Omicron subvariants reduced vaccine effectiveness immediately after the second dose to only about 50% and accelerated the waning of protection.^6–8^ Vaccine protection against Omicron subvariants may not last for more than 6 months after the second dose.^6–8^ Booster vaccination restores vaccine protection to the level observed immediately after the second dose,^8, 9^ but this protection also wanes with time and in a similar pattern to that of the primary series.^6–10^

Though protection of natural infection against reinfection is high immediately after infection,^11, 12^ the protection wanes with time and is not expected to last for more than 3 years.^13^ Introduction of Omicron subvariants reduced pre-Omicron infection protection against Omicron reinfection to only about 50%,^14, 15^ and accelerated the waning of this protection.^13^ Protection of a pre-Omicron infection against Omicron reinfection may not last for more than 1 year.^13, 16^ While protection of Omicron infection against Omicron reinfection is high,^16–18^ evidence suggests that this protection is declining fairly rapidly due to combined effect of waning of protection over time and progressive immune evasiveness of Omicron subvariants.^16^

These immune protection patterns demonstrate the need for the concept of *population immunity* to track evolution of population-level immune protection over time in a given population. Population immunity of primary-series vaccination is defined as the overall protection against infection at a given point in time among persons with only primary-series vaccination relative to those unvaccinated. Booster and previous-infection population immunities are defined analogously. These three forms of population immunity are expected to vary fairly rapidly even within a time horizon of only 1 year due to the combined effects of waning of vaccine and natural immunity and viral immune evasion. Such rapid variation may create fertile grounds for repeated waves of infection to occur even within few months of each other.

We aimed to characterize variation of these three forms of population immunity over time, month by month, in Qatar, a country that experienced SARS-CoV-2 waves dominated sequentially by the index virus,^19^ Alpha,^20^ Beta,^21^ Omicron BA.1 and BA.2,^14^ Omicron BA.4 and BA.5,^18^ Omicron BA.2.75*,^16^ and currently Omicron XBB*, in addition to a prolonged low-incidence phase dominated by Delta^22^ (Figure 1).

**Figure 1:**
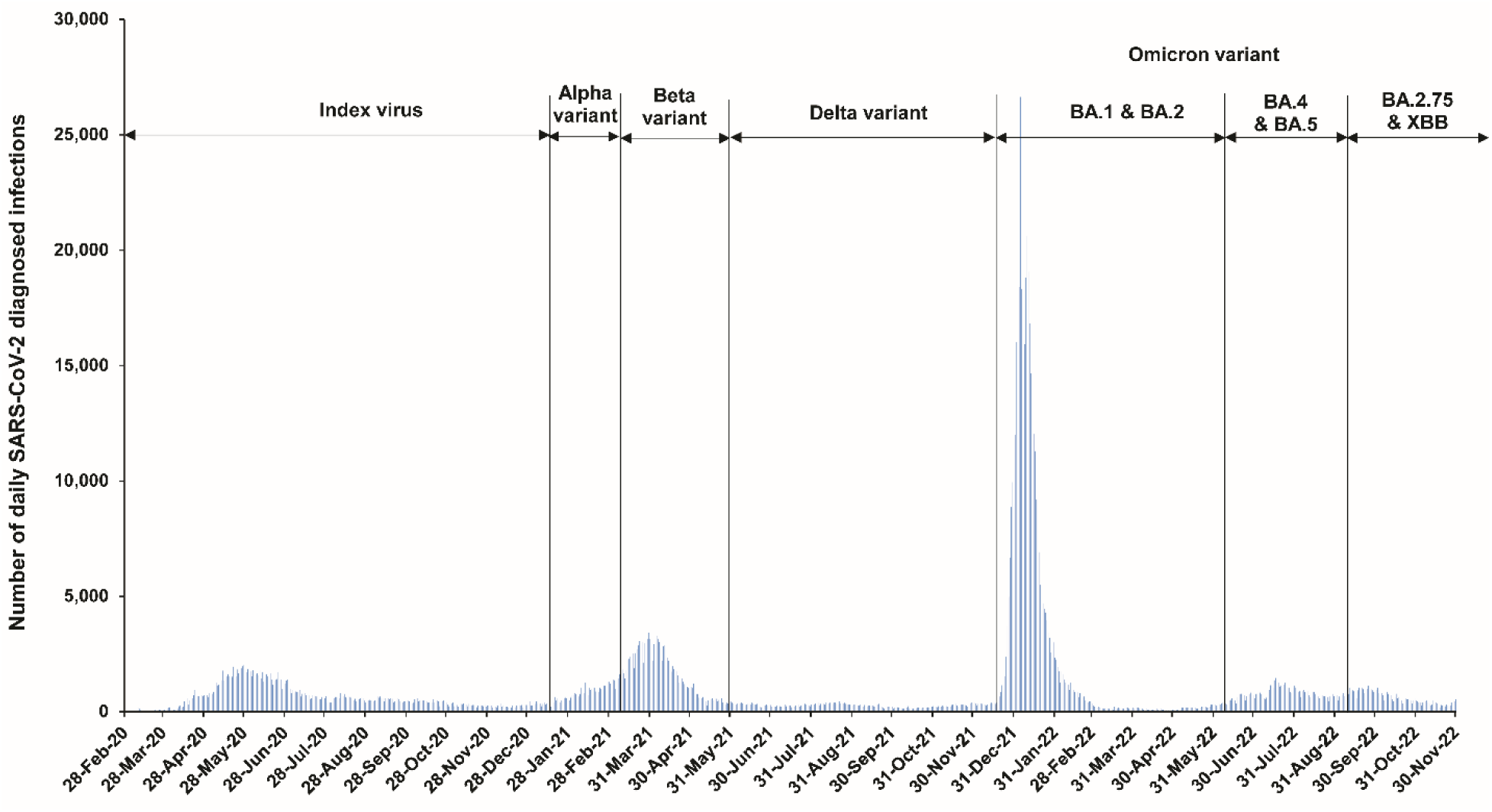
Incidence of SARS-CoV-2 infection in Qatar.

## Methods

### Study population and data sources

This study was conducted on the population of Qatar including data between July 1, 2020 and November 30, 2022. It analyzed the national, federated databases for COVID-19 laboratory testing, vaccination, hospitalization, and death, retrieved from the integrated, nationwide, digital-health information platform (Section S1 of the Supplementary Appendix). Databases include all SARS-CoV-2-related data with no missing information since the onset of the pandemic, including all polymerase chain reaction (PCR) tests, and from January 5, 2022 onward, all medically supervised rapid antigen tests. SARS-CoV-2 testing in Qatar was done at large scale, and up to October 31, 2022, was mostly done for routine reasons such as for screening or travel-related purposes, with infections primarily diagnosed not because of appearance of symptoms, but because of routine testing.^3, 14^ Qatar launched its COVID-19 vaccination program in December of 2020 using almost exclusively the BNT162b2 and mRNA-1273 mRNAvaccines.^23^ Detailed descriptions of Qatar’s population and of the national databases have been reported previously.^3, 9, 10, 14, 19, 24^

### Study design

Effectiveness of previous infection, two-dose (primary-series) vaccination, and third-dose (booster) vaccination in preventing infection, regardless of symptoms, were estimated for Qatar’s population by calendar month. The calendar time over which these effectiveness measures were estimated depended on availability of primary-series and booster vaccination in the population and on having sufficient case numbers to generate estimates.

Each of the effectiveness measures was estimated using the test-negative, case-control, study design, a standard design for assessing immune protection of natural infection^14, 15, 25^ and vaccination.^3, 26–28^ In this design, effectiveness estimates are derived by comparing odds of previous infection or vaccination among positive tests (cases) compared to negative tests (controls).^25–27^

Effectiveness of previous infection in preventing reinfection was defined as the proportional reduction in susceptibility to infection among those with a previous infection versus those without.^12, 14, 25^ Effectiveness of primary-series (or booster) vaccination in preventing infection was defined as the proportional reduction in susceptibility to infection among those vaccinated versus those unvaccinated.^3, 26–28^

All cases (SARS-CoV-2-positive tests) and controls (SARS-CoV-2-negative tests) identified in Qatar during each calendar month of the study duration were matched exactly one-to-one by sex, 10-year age group, nationality, number of coexisting conditions (none, 1, 2, or ≥3), method of testing (PCR or rapid antigen), reason for SARS-CoV-2 testing, and (by design) calendar month of testing. Additionally, cases and controls were matched by COVID-19 vaccine type and number of vaccine doses at time of SARS-CoV-2 test in all analyses of effectiveness of previous infection, and by status of most recent prior infection (no documented prior infection, documented pre-Omicron prior infection, or documented Omicron prior infection) in all vaccine effectiveness analyses.

Matching was done to balance observed confounders between exposure groups that are related to risk of infection in Qatar.^19, 29–32^ Matching by the considered factors was informed by results of prior studies that used matching to control for differences in infection risk in Qatar, including test-negative, case-control studies.^2–4, 23, 33^ For each calendar month, only the first SARS-CoV-2-positive test for each case and the first SARS-CoV-2-negative test for each control were included. Persons qualified as controls if they had at least one SARS-CoV-2-negative test and no record of a SARS-CoV-2-positive test in the considered calendar month.

All persons who received a vaccine other than BNT162b2 or mRNA-1273, or who received a different mix of vaccines, were excluded. Individuals who received a fourth vaccine dose (second booster dose) prior to the study SARS-CoV-2 test were excluded. Bivalent boosters have only been recently introduced in Qatar and remain at low coverage, thus they were not included in this study. Tests occurring within 14 days of a second dose or 7 days of a third dose were excluded. SARS-CoV-2 tests conducted because of travel-related requirements were excluded.^3, 4^ These inclusion and exclusion criteria were implemented to allow for build-up of immunity after vaccination,^1, 9^ and to minimize different types of potential bias, as informed by earlier analyses on the same population.^3, 4^ Every case (or control) that met the inclusion criteria and that could be matched to a control (case) was included in the analysis. Previous infection status and COVID-19 vaccination status were ascertained at the time of the SARS-CoV-2 test.

SARS-CoV-2 reinfection is conventionally defined as a documented infection ≥90 days after an earlier infection, to avoid misclassification of prolonged SARS-CoV-2 positivity as reinfection, if a shorter time interval is used.^34, 35^ Previous infection was thus defined as a SARS-CoV-2-positive test ≥90 days before the study’s SARS-CoV-2 test.^14, 15, 25^ Cases or controls with SARS-CoV-2-positive tests <90 days before the study’s SARS-CoV-2 test were excluded.

Effectiveness was also estimated against severe, critical, or fatal COVID-19 due to SARS-CoV-2 infection using the same methodology, but for 6-calendar-month durations owing to the small number of cases with severe forms of COVID-19. Cases and controls were matched one-to-five to increase precision of estimates. Classification of COVID-19 case severity (acute-care hospitalizations),^36^ criticality (intensive-care-unit hospitalizations),^36^ and fatality^37^ followed World Health Organization (WHO) guidelines, and assessments were made by trained medical personnel using individual chart reviews (Section S3).

Each person who had a SARS-CoV-2-positive test and COVID-19 hospital admission was subject to an infection severity assessment every three days until discharge or death, regardless of the length of hospital stay or the time between the SARS-CoV-2-positive test and the final disease outcome. Individuals who progressed to severe,^36^ critical,^36^ or fatal^37^ COVID-19 between the SARS-CoV-2-positive test and the end of study were classified based on their worst outcome, starting with death, followed by critical disease, and then severe disease.

### Oversight

The institutional review boards at Hamad Medical Corporation and Weill Cornell Medicine– Qatar approved this retrospective study with a waiver of informed consent. The study was reported according to the Strengthening the Reporting of Observational Studies in Epidemiology (STROBE) guidelines (Table S1).

### Statistical analysis

While all records of SARS-CoV-2 testing were examined for selection of cases and controls, only matched samples were analyzed. Cases and controls were described using frequency distributions and measures of central tendency and compared using standardized mean differences (SMDs). An SMD of ≤0.1 indicated adequate matching.^38^ The median and interquartile range (IQR) of the duration between the immunological event (previous infection, primary-series vaccination, or booster vaccination) and SARS-CoV-2 test were calculated for cases and controls in each calendar-month analysis.

The odds ratio (and its associated 95% confidence interval (CI)), comparing odds of the immunological event among cases to that among controls, was estimated using conditional logistic regression, that is factoring the matching in the study design. This analytical approach was implemented to reduce potential bias due to variation in epidemic phase,^26, 39^ gradual vaccination roll-out,^26, 39^ and other confounders.^19, 40, 41^ CIs did not factor multiplicity and interactions were not examined.

Vaccine effectiveness and associated 95% CI were estimated as 1-odds ratio (OR) of the immunological event among cases versus controls if the OR was <1,^25–27^ and as (1/OR)-1 if the OR was ≥1.^10, 42^ The latter was done to ensure symmetric scale for both negative and positive effectiveness, ranging from -100%-100%.^10, 42^ Statistical analyses were conducted in STATA/SE version 17.0 (Stata Corporation, College Station, TX, USA).

## Results

### Effectiveness of previous infection against reinfection

Characteristics of matched cases and controls of the study population are shown in Table S2. Effectiveness of previous infection against reinfection was high from July of 2020 to November of 2021, a duration coinciding with incidence of pre-Omicron variants (Figure 1), at a level that exceeded 70% (Table S3 and Figure 2A). Effectiveness declined slowly over time after a wave, but rebounded to a higher level after a new wave. Note that since cases and controls with SARS-CoV-2-positive tests <90 days before the study’s SARS-CoV-2 test were excluded to avoid misclassification of prolonged SARS-CoV-2 positivity as reinfection,^34, 35^ estimates of the effectiveness of previous infections represent infections that occurred ≥90 days prior and not that of recent infections.

**Figure 2:**
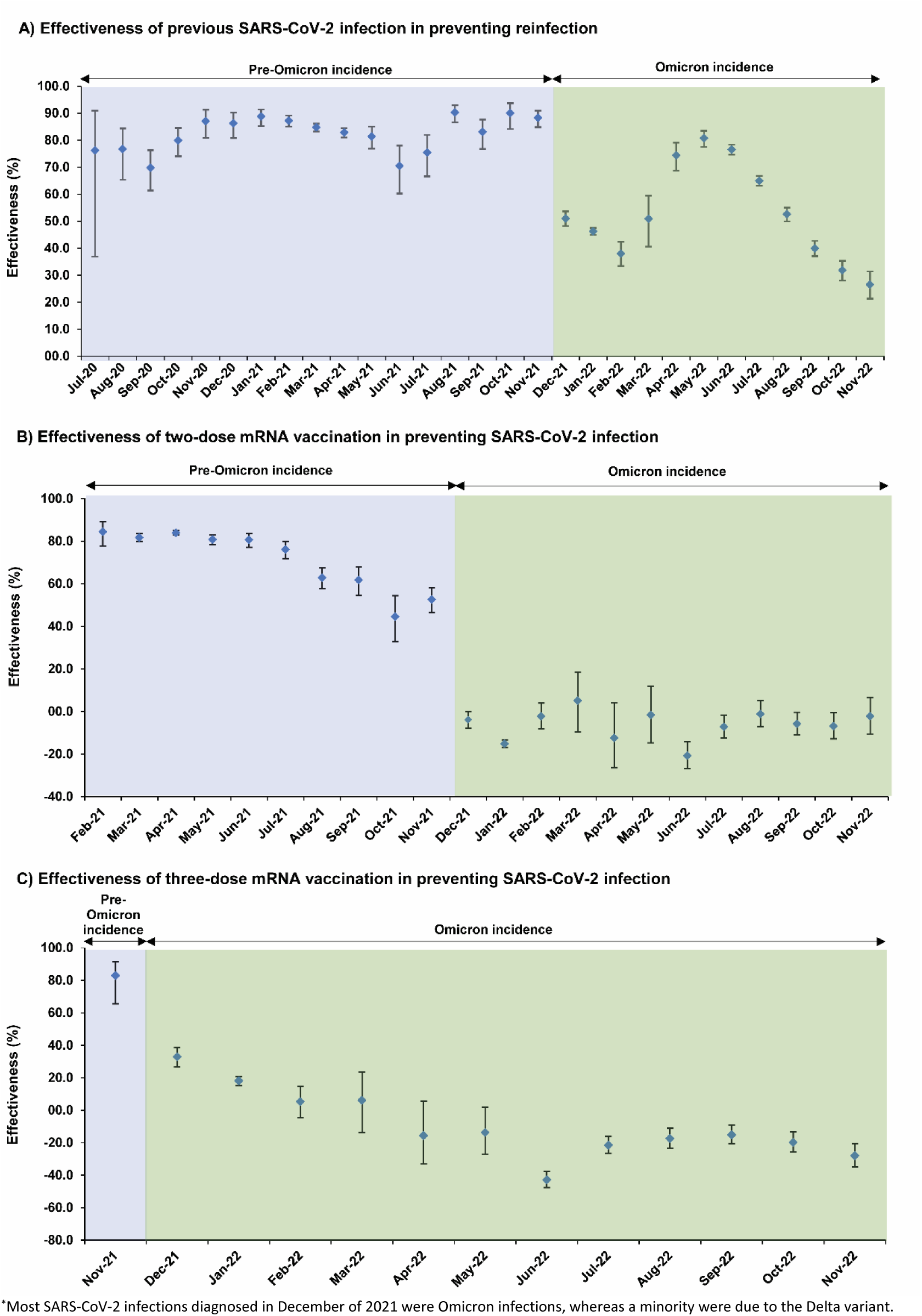
Effectiveness of A) previous SARS-CoV-2 infection in preventing reinfection, B) primary-series (two-dose) mRNA vaccination in preventing infection, and C) booster (third-dose) mRNA vaccination in preventing infection, in Qatar, between July of 2020 and November of 2022*.

Effectiveness dropped immediately and immensely with the emergence of Omicron in December 2021 (Table S3 and Figure 2A). While effectiveness was 88.3% (95% CI: 84.8-91.0%) in November 2021, it dropped to 51.0% (95% CI: 48.3-53.6%) in December 2021; a drop of ∼40 absolute percentage points within one month associated with the change in the circulating variant. Effectiveness continued at this level during the large BA.1/BA.2 Omicron wave, up to March 2022.

Effectiveness rebounded again after end of the BA.1/BA.2 wave and reached 80.8% (95% CI: 77.6-83.5%) in May of 2022 before gradually declining in subsequent months (Table S3 and Figure 2A). Effectiveness was low at ∼30% by end of the study in November of 2022, a time coinciding with incidence of BA.2.75* and XBB* subvariants (Figure 1).

### Effectiveness of primary-series vaccination against infection

Characteristics of matched cases and controls of the study population are shown in Table S2. Effectiveness of primary-series vaccination against infection was very high at >80% right after introduction of vaccination (Table S4 and Figure 2B). Effectiveness was 84.5% (95% CI: 77.8-89.2%) in February of 2021 and remained high up to June of 2021, coinciding with the rapid scale-up of vaccination in Qatar^3^ and incidence of pre-Omicron variants (Figure 1).

However, starting from July of 2021, when the mass vaccination campaigns slowed down along with a decline in the recentness of the second dose for much of the population, effectiveness started to decline rapidly (Table S4 and Figure 2B). Effectiveness was 80.7% (95% CI: 77.2-83.7%) in June of 2021 before dropping to 76.2% (95% CI: 71.8-79.9%) in July of 2021, 63.0% (95% CI: 57.8-67.5%) in August of 2021, 61.9% (95% CI: 54.7-68.0%) in September of 2021, 44.6% (95% CI: 32.8-54.4%) in October of 2021, and 52.7% (95% CI: 46.5-58.2%) in November of 2021, right before introduction of Omicron (Figure 1).

Effectiveness declined linearly with calendar time by ∼1 absolute percentage point every 5 days (Figure 3A). Effectiveness declined linearly with median time since the second dose also by ∼1 absolute percentage point every 5 days (Figure 3B). Extrapolating this linear trend indicated that effectiveness would reach 0% in 13.4 months after the second dose.

**Figure 3:**
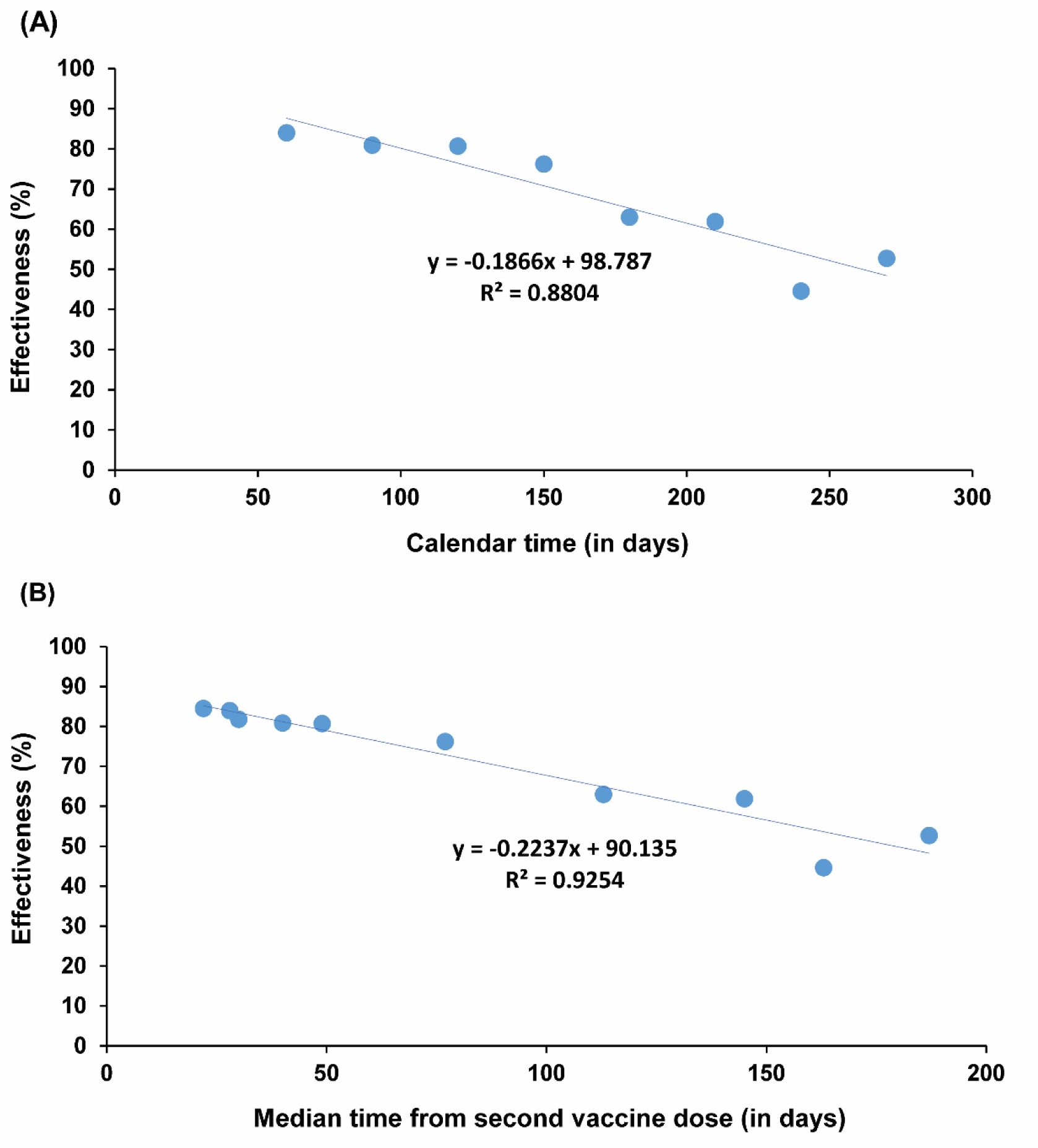
Association between the effectiveness (per calendar month) of primary-series mRNA vaccination in preventing infection and (A) calendar time (in days) and (B) median time from second vaccine dose (in days).

Effectiveness dropped suddenly and massively with introduction of Omicron in December of 2021 (Table S4 and Figure 2B). While effectiveness was 52.7% (95% CI: 46.5-58.2%) in November of 2021, it became negligible during the BA.1/BA.2 wave starting from December of 2021, that is a drop of ∼50 absolute percentage points within one month associated with the change in circulating variant. Effectiveness remained negligible in subsequent months and was notably negative during the BA.4/BA.5 wave in June and July of 2022.

### Effectiveness of booster vaccination against infection

Characteristics of matched cases and controls of the study population are shown in Table S2. Effectiveness of booster (third dose) vaccination against infection was very high at 83.0% (95% CI: 65.6-91.6%) in November of 2021 (Table S5 and Figure 2C), when booster vaccination was being scaled up^9^ and Delta was circulating (Figure 1). But effectiveness dropped suddenly and immensely with the emergence of Omicron in December of 2021. Effectiveness dropped from 83.0% (95% CI: 65.6-91.6%) in November of 2021 to 32.9% (95% CI: 26.7-38.5%) in December of 2021, that is a drop of ∼50 absolute percentage points within one month associated with the change in the circulating variant. Effectiveness continued to decline thereafter and became negative between April and November of 2022, particularly during the BA.4/BA.5 wave.

### Effectiveness against severe, critical, or fatal COVID-19

Unlike effectiveness against infection, effectiveness against severe, critical, or fatal COVID-19 of previous infection (Table S6 and Figure 4A), primary-series vaccination (Table S6 and Figure 4B), and booster vaccination (Table S6 and Figure 4C) were high throughout the study duration at generally >80%. There appeared to be some waning in effectiveness of primary-series vaccination and booster vaccination over time, but the number of severe COVID-19 cases was too small to allow precise estimation of potential waning in effectiveness.

**Figure 4:**
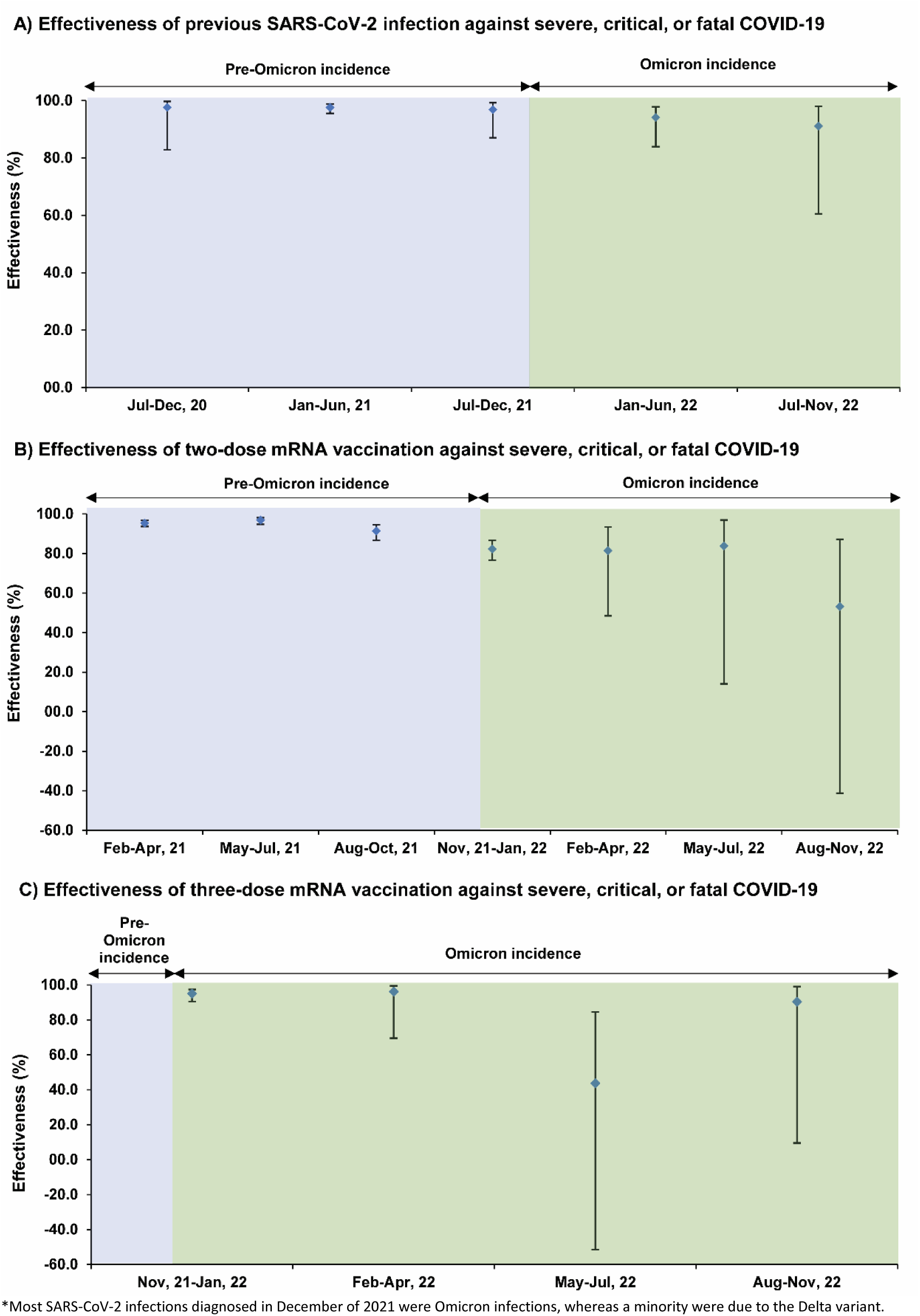
Effectiveness against severe, critical, or fatal COVID-19 of A) previous SARS-CoV-2 infection, B) primary-series (two-dose) mRNA vaccination, and C) booster (three-dose) mRNA vaccination, in Qatar, between July of 2020 and November of 2022*.

## Discussion

This study investigated three population immunity patterns of relevance to the future of this pandemic. First, both natural and vaccine population immunity wane with time, with the waning being particularly rapid for vaccine population immunity. Even before Omicron introduction, vaccine protection was waning by ∼1 absolute percentage point every 5 days. The protection induced by vaccination appeared to fully wane within 14 months. These findings suggest that repeated waves of infection will become a generic feature of SARS-CoV-2 epidemiology. High levels of population immunity may not be sustained for more than a year or so.

Second, sudden emergence of a new variant that is substantially different from previously circulating variants can immensely and immediately affect population immunity. Protection of previous infection dropped by ∼40 absolute percentage points within only one month after Omicron introduction. Similarly, each of primary-series and booster protections dropped by ∼50 absolute percentage points. As a result, Qatar experienced its largest SARS-CoV-2 epidemic wave to date (Figure 1). This demonstrates how the long-term epidemiology of this infection can be strongly sensitive to major shifts in virus evolution. Active surveillance of emerging variants and development of an early warning system are critical to mitigate the consequences of immune-evasive variants.^43^

Third, although population immunity against infection waned rapidly, population immunity against severe COVID-19 was durable and showed slow waning even with introduction of Omicron. Infection with common-cold coronaviruses,^44^ and perhaps influenza,^45^ induces only a year-long protection against infection, but life-long immunity against severe reinfection.^46^ While it is premature to make long-term predictions, this finding suggests that SARS-CoV-2 epidemiology may exhibit a similar pattern to that of common-cold coronaviruses. Long-term immune protection against severe COVID-19 could contribute to a benign pattern of infection that is perhaps not dissimilar to that of common-cold coronaviruses.

Some of the vaccine effectiveness measures post Omicron introduction, particularly for booster vaccination, were negative in value, perhaps suggesting negative immune imprinting. This effect was pronounced during the BA.4/BA.5 wave. This finding supports similar recent findings in this same population.^10, 47^ Such imprinting effects have been observed for other infections such as influenza.^48, 49^ It remains to be seen whether these effects will be of consequence in the future epidemiology of SARS-CoV-2 infection.

This study has limitations. Since SARS-CoV-2 reinfection is defined as an infection ≥90 days after an earlier infection, estimates of previous-infection protection lagged actual level of protection by 3 months. This has underestimated previous-infection population immunity during or right after large waves, when population immunity was building up rapidly. With the relatively young population of Qatar, our findings may not be generalizable to other countries where elderly citizens constitute a larger proportion of the total population.

At this stage of the pandemic, the reference groups of persons with no previous infection and unvaccinated persons may not be representative due to depletion of susceptibles and undocumented infections and vaccinations. Vaccinations outside of Qatar may not have been registered in the national database after easing vaccine requirements in 2021. Bias due to depletion of susceptibles may lead to underestimation of previous-infection or vaccine protections,^50^ even in the test-negative study design which is less prone to effect of this bias.^3, 25^ Misclassification of previous-infection status can lead to underestimation of previous-infection effectiveness particularly if the majority of the population had already been infected.^25^ These sources of bias may explain some of the negative vaccine effectiveness values observed in this study post Omicron introduction and the lower than expected previous-infection protection in the last few months of the study. Estimates after the first large Omicron wave may provide qualitative findings rather than precise quantitative estimates.

Bias can arise in real-world observational data in unexpected ways, or from unknown sources, such as subtle differences in test-seeking behavior or changes in the pattern of testing due to policy changes, tests’ accessibility, or behavioral differences (Section S1). The reason for testing varies with time even if there are no policy changes. During a wave, a large proportion of testing occurs because of clinical suspicion of infection. At times of low incidence, a large proportion of testing occurs because of routine reasons.

The study did not investigate population immunity of a second booster dose or of bivalent boosters due to the low coverage of these vaccinations in Qatar. In the first few weeks of the pandemic, PCR workflows were still under development and there was a chance of false-positive tests. While this may have affected only a very small number of PCR tests, reinfections were so rare at this time that few such false-positive cases could have resulted in underestimation of previous-infection protection in the first few months of the pandemic.

While matching was done for several factors, it was not possible for other factors such as geography or occupation, as such data were unavailable. However, Qatar is essentially a city state and infection incidence was broadly distributed across neighborhoods. Nationality, age, and sex provide a powerful proxy for socio-economic status in this country,^19, 29–32^ and thus matching by these factors may have partially controlled for other factors such as occupation. The matching prescription had already been investigated in previous studies of different epidemiologic designs, and using control groups to test for null effects.^2–4, 23, 33^ These studies have supported that this prescription provides adequate control of the differences in infection exposure.^2–4, 23, 33^ The study was implemented on Qatar’s total population, perhaps minimizing the likelihood of bias.

In conclusion, both previous-infection and vaccine population immunities wane with time and the waning is rapid for vaccine protection. High levels of population immunity may not be sustained for more than a year. The waning of population immunity facilitates fertile grounds for repeated waves of infection to occur even within few months of each other. Timely administration of boosters for those vulnerable to COVID-19 may remain essential for years to come. Emergence of a new variant that is substantially different from previously circulating variants can suddenly and immensely reduce population immunity leading to large epidemic waves. Repeated waves of infection may also facilitate further evolution of the virus and continual immune evasion. However, with the durability of population immunity against severe COVID-19, the repeated epidemic waves may increasingly exhibit an overall benign pattern of infection.

## Data Availability

The dataset of this study is a property of the Qatar Ministry of Public Health that was provided to the researchers through a restricted-access agreement that prevents sharing the dataset with a third party or publicly. The data are available under restricted access for preservation of confidentiality of patient data. Access can be obtained through a direct application for data access to Her Excellency the Minister of Public Health (https://www.moph.gov.qa/english/OurServices/eservices/Pages/Governmental-HealthCommunication-Center.aspx). The raw data are protected and are not available due to data privacy laws. Aggregate data are available within the paper and its supplementary information.

## Contributors

SHQ, HC, and LJA co-designed the study. SHQ performed the statistical analyses and co-wrote the first draft of the article. LJA conceived the study, led the statistical analyses, and co-wrote the first draft of the article. PVC designed mass PCR testing to allow routine capture of SGTF variants and conducted viral genome sequencing. PT and MRH designed and conducted multiplex, RT-qPCR variant screening and viral genome sequencing. HY, AAA, and HAK conducted viral genome sequencing. All authors contributed to data collection and acquisition, database development, discussion and interpretation of the results, and to the writing of the manuscript. All authors have read and approved the final manuscript.

## Acknowledgements

We acknowledge the many dedicated individuals at Hamad Medical Corporation, the Ministry of Public Health, the Primary Health Care Corporation, Qatar Biobank, Sidra Medicine, and Weill Cornell Medicine-Qatar for their diligent efforts and contributions to make this study possible. The authors are grateful for institutional salary support from the Biomedical Research Program and the Biostatistics, Epidemiology, and Biomathematics Research Core, both at Weill Cornell Medicine-Qatar, as well as for institutional salary support provided by the Ministry of Public Health, Hamad Medical Corporation, and Sidra Medicine. The authors are also grateful for the Qatar Genome Programme and Qatar University Biomedical Research Center for institutional support for the reagents needed for the viral genome sequencing. HHA acknowledges the support of Qatar University Internal Grant ID QUCG-CAS-23/24-114. The funders of the study had no role in study design, data collection, data analysis, data interpretation, or writing of the article. Statements made herein are solely the responsibility of the authors.

## Supplementary Appendix

### *Section S1*: Further details on methods

#### Data sources and testing

Qatar’s national and universal public healthcare system uses the Cerner-system advanced digital health platform to track all electronic health record encounters of each individual in the country, including all citizens and residents registered in the national and universal public healthcare system. Registration in the public healthcare system is mandatory for citizens and residents.

The databases analyzed in this study are data-extract downloads from the Cerner-system that have been implemented on a regular (twice weekly) schedule since the onset of the pandemic by the Business Intelligence Unit at Hamad Medical Corporation. Hamad Medical Corporation (HMC) is the national public healthcare provider in Qatar. At every download all tests, coronavirus disease 2019 (COVID-19) vaccinations, hospitalizations related to COVID-19, and all death records regardless of cause are provided to the authors through .csv files. These databases have been analyzed throughout the pandemic not only for study-related purposes, but also to provide policymakers with summary data and analytics to inform the national response.

Every health encounter in the Cerner-system is linked to a unique individual through the HMC Number that links all records for this individual at the national level. Databases were merged and analyzed using the HMC Number to link all records whether for testing, vaccinations, hospitalizations, and deaths. All deaths in Qatar are tracked by the public healthcare system. All COVID-19-related healthcare was provided only in the public healthcare system. No private entity was permitted to provide COVID-19-related healthcare. COVID-19 vaccination was also provided only through the public healthcare system. These health records were tracked throughout the COVID-19 pandemic using the Cerner system. This system has been implemented in 2013, before the onset of the pandemic.

Demographic details for every HMC Number (individual) such as sex, age, and nationality are collected upon issuing of the universal health card, based on the Qatar Identity Card, which is a mandatory requirement by the Ministry of Interior to every citizen and resident in the country.

Severe acute respiratory syndrome coronavirus 2 (SARS-CoV-2) testing in the healthcare system in Qatar is done at a mass scale, and up to October 31, 2022, was mostly done for routine reasons, where about 5% of the population were tested every week.^1, 2^ All SARS-CoV-2 testing in any facility in this country is tracked nationally in one database, the national testing database. This database covers all testing in all locations and facilities throughout the country, whether public or private. Every polymerase chain reaction (PCR) test and an increasing proportion of the medically supervised rapid antigen tests conducted in Qatar, regardless of location or setting, are classified on the basis of symptoms and the reason for testing (clinical symptoms, contact tracing, surveys or random testing campaigns, individual requests, routine healthcare testing, pre-travel, at port of entry, or other). Based on the distribution of the reason for testing up to October 31, 2022, most of the tests that have been conducted in Qatar were conducted for routine reasons, such as being travel-related. About 75% of those diagnosed are also diagnosed not because of appearance of symptoms, but because of routine testing.^1, 2^

The first large Omicron wave that peaked in January of 2022 was massive and strained the testing capacity in the country.^1, 3^ Accordingly, rapid antigen testing was introduced to relieve the pressure on PCR testing. Implementation of this change in testing occurred quickly precluding incorporation of reason for testing in a large proportion of the rapid antigen tests for several months. While the reason for testing is available for all PCR tests, it is not available for all rapid antigen tests. Availability of reason for testing for the rapid antigen tests also varied with time. Rapid antigen test kits are available for purchase in pharmacies in Qatar, but outcome of home-based testing is not reported nor documented in the national databases. Since SARS-CoV-2-test outcomes are linked to specific public health measures, restrictions, and privileges, testing policy and guidelines stress facility-based testing as the core testing mechanism in the population. While facility-based testing is provided free of charge or at low subsidized costs, depending on the reason for testing, home-based rapid antigen testing is de-emphasized and not supported as part of national policy.

Qatar has unusually young, diverse demographics, in that only 9% of its residents are ≥50 years of age, and 89% are expatriates from over 150 countries.^4, 5^ Further descriptions of the study population and these national databases were reported previously.^1, 2, 5–7^

#### Comorbidity classification

Comorbidities were ascertained and classified based on the ICD-10 codes as recorded in the electronic health record encounters of each individual in the Cerner-system national database that includes all citizens and residents registered in the national and universal public healthcare system. The public healthcare system provides healthcare to the entire resident population of Qatar free of charge or at heavily subsidized costs, including prescription drugs.

All encounters for each individual were analyzed to determine the comorbidity classification for that individual, as part of a recent national analysis to assess healthcare needs and resource allocation. The Cerner-system national database includes encounters starting from 2013, after this system was launched in Qatar. As long as each individual had at least one encounter with a specific comorbidity diagnosis since 2013, this person was classified with this comorbidity. Individuals who have comorbidities but never sought care in the public healthcare system, or seek care exclusively in private healthcare facilities, were classified as individuals with no comorbidity due to absence of recorded encounters for them.

### *Section S2*: Laboratory methods and variant ascertainment

#### Real-time reverse-transcription polymerase chain reaction testing

Nasopharyngeal and/or oropharyngeal swabs were collected for polymerase chain reaction (PCR) testing and placed in Universal Transport Medium (UTM). Aliquots of UTM were: 1) extracted on KingFisher Flex (Thermo Fisher Scientific, USA), MGISP-960 (MGI, China), or ExiPrep 96 Lite (Bioneer, South Korea) followed by testing with real-time reverse-transcription PCR (RT-qPCR) using TaqPath COVID-19 Combo Kits (Thermo Fisher Scientific, USA) on an ABI 7500 FAST (Thermo Fisher Scientific, USA); 2) tested directly on the Cepheid GeneXpert system using the Xpert Xpress SARS-CoV-2 (Cepheid, USA); or 3) loaded directly into a Roche cobas 6800 system and assayed with the cobas SARS-CoV-2 Test (Roche, Switzerland). The first assay targets the viral S, N, and ORF1ab gene regions. The second targets the viral N and E-gene regions, and the third targets the ORF1ab and E-gene regions.

All PCR testing was conducted at the Hamad Medical Corporation Central Laboratory or Sidra Medicine Laboratory, following standardized protocols.

#### Rapid antigen testing

SARS-CoV-2 antigen tests were performed on nasopharyngeal swabs using one of the following lateral flow antigen tests: Panbio COVID-19 Ag Rapid Test Device (Abbott, USA); SARS-CoV-2 Rapid Antigen Test (Roche, Switzerland); Standard Q COVID-19 Antigen Test (SD Biosensor, Korea); or CareStart COVID-19 Antigen Test (Access Bio, USA). All antigen tests were performed point-of-care according to each manufacturer’s instructions at public or private hospitals and clinics throughout Qatar with prior authorization and training by the Ministry of Public Health (MOPH). Antigen test results were electronically reported to the MOPH in real time using the Antigen Test Management System which is integrated with the national Coronavirus Disease 2019 (COVID-19) database.

#### Classification of infections by variant type

Surveillance for SARS-CoV-2 variants in Qatar is based on viral genome sequencing and multiplex RT-qPCR variant screening^8^ of random positive clinical samples,^2, 9–13^ complemented by deep sequencing of wastewater samples.^11, 14, 15^ Further details on the viral genome sequencing and multiplex RT-qPCR variant screening throughout the SARS-CoV-2 waves in Qatar can be found in previous publications.^1–3, 7, 9–13, 16–20^

### *Section S3*: COVID-19 severity, criticality, and fatality classification

Classification of COVID-19 case severity (acute-care hospitalizations),^21^ criticality (intensive-care-unit hospitalizations),^21^ and fatality^22^ followed World Health Organization (WHO) guidelines. Assessments were made by trained medical personnel independent of study investigators and using individual chart reviews, as part of a national protocol applied to every hospitalized COVID-19 patient. Each hospitalized COVID-19 patient underwent an infection severity assessment every three days until discharge or death. We classified individuals who progressed to severe, critical, or fatal COVID-19 between the time of the documented infection and the end of the study based on their worst outcome, starting with death,^22^ followed by critical disease,^21^ and then severe disease.^21^

Severe COVID-19 disease was defined per WHO classification as a SARS-CoV-2 infected person with “oxygen saturation of <90% on room air, and/or respiratory rate of >30 breaths/minute in adults and children >5 years old (or ≥60 breaths/minute in children <2 months old or ≥50 breaths/minute in children 2-11 months old or ≥40 breaths/minute in children 1–5 years old), and/or signs of severe respiratory distress (accessory muscle use and inability to complete full sentences, and, in children, very severe chest wall indrawing, grunting, central cyanosis, or presence of any other general danger signs)”.^21^ Detailed WHO criteria for classifying SARS-CoV-2 infection severity can be found in the WHO technical report.^21^

Critical COVID-19 disease was defined per WHO classification as a SARS-CoV-2 infected person with “acute respiratory distress syndrome, sepsis, septic shock, or other conditions that would normally require the provision of life sustaining therapies such as mechanical ventilation (invasive or non-invasive) or vasopressor therapy”.^21^ Detailed WHO criteria for classifying SARS-CoV-2 infection criticality can be found in the WHO technical report.^21^

COVID-19 death was defined per WHO classification as “a death resulting from a clinically compatible illness, in a probable or confirmed COVID-19 case, unless there is a clear alternative cause of death that cannot be related to COVID-19 disease (e.g. trauma). There should be no period of complete recovery from COVID-19 between illness and death. A death due to COVID-19 may not be attributed to another disease (e.g. cancer) and should be counted independently of preexisting conditions that are suspected of triggering a severe course of COVID-19”. Detailed WHO criteria for classifying COVID-19 death can be found in the WHO technical report.^22^

---

**Table S1:** STROBE checklist for case-control studies.

|  | Item No | Recommendation | Main text page |
| --- | --- | --- | --- |
| Title and abstract | 1 | (a) Indicate the study’s design with a commonly used term in the title or the abstract | Abstract |
|  |  | (b) Provide in the abstract an informative and balanced summary of what was done and what was found | Abstract |
| Introduction |  |  |  |
| Background/rationale | 2 | Explain the scientific background and rationale for the investigation being reported | Introduction |
| Objectives | 3 | State specific objectives, including any prespecified hypotheses | Introduction |
| Methods |  |  |  |
| Study design | 4 | Present key elements of study design | Methods (‘Study design’) |
| Setting | 5 | Describe the setting, locations, and relevant dates, including periods of recruitment, exposure, follow-up, and data collection | Methods (‘Study population and data sources’ & ‘Study design’) & Section S1 in Supplementary Appendix |
| Participants | 6 | (a) Give the eligibility criteria, and the sources and methods of case ascertainment and control selection. Give the rationale for the choice of cases and controls<br>(b) For matched studies, give matching criteria and the number of controls per case | Methods (‘Study design’) |
| Variables | 7 | Clearly define all outcomes, exposures, predictors, potential confounders, and effect modifiers. Give diagnostic criteria, if applicable | Methods (‘Study design’ & ‘Statistical analysis’) & Sections S1-S3 in Supplementary Appendix |
| Data sources/measurement | 8 | For each variable of interest, give sources of data and details of methods of assessment (measurement). Describe comparability of assessment methods if there is more than one group | Methods (‘Study population and data sources’, ‘Study design’ & ‘Statistical analysis’, paragraph 1) & Sections S1-S3 in Supplementary Appendix |
| Bias | 9 | Describe any efforts to address potential sources of bias | Methods (‘Study design’ & ‘Statistical analysis’) |
| Study size | 10 | Explain how the study size was arrived at | Methods (‘Study population and data sources’ & ‘Study design’) |
| Quantitative variables | 11 | Explain how quantitative variables were handled in the analyses. If applicable, describe which groupings were chosen and why | Methods (‘Study design’ & ‘Statistical analysis’) |
| Statistical methods | 12 | (a) Describe all statistical methods, including those used to control for confounding | Methods (‘Statistical analysis’) |
|  |  | (b) Describe any methods used to examine subgroups and interactions | Methods (‘Statistical analysis’) |
|  |  | (c) Explain how missing data were addressed | Not applicable, see Methods (‘Study population and data sources’) |
|  |  | (d) If applicable, explain how matching of cases and controls was addressed | Methods (‘Study design’ & ‘Statistical analysis’) |
|  |  | (e) Describe any sensitivity analyses | Not applicable |
| Results |  |  |  |
| Participants | 13 | (a) Report numbers of individuals at each stage of study—eg numbers potentially eligible, examined for eligibility, confirmed eligible, included in the study, completing follow-up, and analysed<br>(b) Give reasons for non-participation at each stage<br>(c) Consider use of a flow diagram | Tables S3-S6 in Supplementary Appendix |
| Descriptive data | 14 | (a) Give characteristics of study participants (eg demographic, clinical, social) and information on exposures and potential confounders<br>(b) Indicate number of participants with missing data for each variable of interest | Tables S2-S6 in Supplementary Appendix<br>Not applicable, see Methods (‘Study population and data sources’) |
| Outcome data | 15 | Report numbers in each exposure category, or summary measures of exposure | Results, Figures 2 & 4, & Tables S3-S6 in Supplementary Appendix |
| Main results | 16 | (a) Give unadjusted estimates and, if applicable, confounder-adjusted estimates and their precision (eg, 95% confidence interval). Make clear which confounders were adjusted for and why they were included | Results, Figures 2 & 4, & Tables S3-S6 in Supplementary Appendix |
|  |  | (b) Report category boundaries when continuous variables were categorized | Figures 2 & 4, & Tables S3-S6 in Supplementary Appendix |
|  |  | (c) If relevant, consider translating estimates of relative risk into absolute risk for a meaningful time period | Not applicable |
| Other analyses | 17 | Report other analyses done—eg analyses of subgroups and interactions, and sensitivity analyses | Not applicable |
| Discussion |  |  |  |
| Key results | 18 | Summarise key results with reference to study objectives | Discussion, paragraphs 1-4 |
| Limitations | 19 | Discuss limitations of the study, taking into account sources of potential bias or imprecision. Discuss both direction and magnitude of any potential bias | Discussion, paragraphs 5-9 |
| Interpretation | 20 | Give a cautious overall interpretation of results considering objectives, limitations, multiplicity of analyses, results from similar studies, and other relevant evidence | Discussion, paragraph 10 |
| Generalisability | 21 | Discuss the generalisability (external validity) of the study results | Discussion, paragraphs 5-6 |
| **Other information** |  |  |  |
| Funding | 22 | Give the source of funding and the role of the funders for the present study and, if applicable, for the original study on which the present article is based | Acknowledgements |

**Table S2:** Characteristics of matched cases and controls using the combined samples over all months of the studies investigating effectiveness against SARS-CoV-2 infection of previous SARS-CoV-2 infection, primary-series (two-dose) mRNA vaccination, and booster (third dose) mRNA vaccination.

| Characteristics | Study 1<br>Effectiveness of previous SARS-CoV-2 infection |  |  | Study 2<br>Effectiveness of primary-series vaccination |  |  | Study 3<br>Effectiveness of booster vaccination |  |  |
| --- | --- | --- | --- | --- | --- | --- | --- | --- | --- |
|  | Cases*<br>n (%) | Controls*<br>n (%) | SMD<sup>†</sup> | Cases<sup>‡</sup><br>n (%) | Controls<sup>‡</sup><br>n (%) | SMD<sup>†</sup> | Cases<sup>‡</sup><br>n (%) | Controls<sup>‡</sup><br>n (%) | SMD<sup>†</sup> |
|  | N=530,213 | N=530,213 |  | N=382,978 | N=382,978 |  | N=156,115 | N=156,115 |  |
| Median age (IQR)—years | 32 (20-40) | 32 (20-40) | 0.00<sup>§</sup> | 30 (15-39) | 30 (16-39) | 0.00<sup>§</sup> | 27 (8-38) | 27 (8-38) | 0.00<sup>§</sup> |
| Age—years |  |  |  |  |  |  |  |  |  |
| 0-9 years | 70,051 (13.2) | 70,051 (13.2) |  | 60,438 (15.8) | 60,438 (15.8) |  | 45,058 (28.9) | 45,058 (28.9) |  |
| 10-19 years | 62,049 (11.7) | 62,049 (11.7) |  | 51,444 (13.4) | 51,444 (13.4) |  | 19,896 (12.7) | 19,896 (12.7) |  |
| 20-29 years | 98,917 (18.7) | 98,917 (18.7) |  | 73,603 (19.2) | 73,603 (19.2) |  | 20,106 (12.9) | 20,106 (12.9) |  |
| 30-39 years | 160,078 (30.2) | 160,078 (30.2) |  | 110,005 (28.7) | 110,005 (28.7) |  | 35,226 (22.6) | 35,226 (22.6) |  |
| 40-49 years | 90,418 (17.1) | 90,418 (17.1) | 0.00 | 58,939 (15.4) | 58,939 (15.4) | 0.00 | 21,853 (14.0) | 21,853 (14.0) | 0.00 |
| 50-59 years | 34,878 (6.6) | 34,878 (6.6) |  | 20,557 (5.4) | 20,557 (5.4) |  | 9,483 (6.1) | 9,483 (6.1) |  |
| 60-69 years | 10,369 (2.0) | 10,369 (2.0) |  | 5,890 (1.5) | 5,890 (1.5) |  | 3,224 (2.1) | 3,224 (2.1) |  |
| 70-79 years | 2,558 (0.5) | 2,558 (0.5) |  | 1,522 (0.4) | 1,522 (0.4) |  | 914 (0.6) | 914 (0.6) |  |
| 80+ years | 895 (0.2) | 895 (0.2) |  | 580 (0.2) | 580 (0.2) |  | 355 (0.2) | 355 (0.2) |  |
| Sex |  |  |  |  |  |  |  |  |  |
| Male | 310,962 (58.7) | 310,962 (58.7) |  | 219,871 (57.4) | 219,871 (57.4) |  | 80,576 (51.6) | 80,576 (51.6) |  |
| Female | 219,251 (41.4) | 219,251 (41.4) | 0.00 | 163,107 (42.6) | 163,107 (42.6) | 0.00 | 75,539 (48.4) | 75,539 (48.4) | 0.00 |
| Nationality<sup>¶</sup> |  |  |  |  |  |  |  |  |  |
| Bangladeshi | 25,361 (4.8) | 25,361 (4.8) |  | 17,700 (4.6) | 17,700 (4.6) |  | 3,029 (1.9) | 3,029 (1.9) |  |
| Egyptian | 32,437 (6.1) | 32,437 (6.1) |  | 23,224 (6.1) | 23,224 (6.1) |  | 10,584 (6.8) | 10,584 (6.8) |  |
| Filipino | 57,347 (10.8) | 57,347 (10.8) |  | 37,274 (9.7) | 37,274 (9.7) |  | 18,211 (11.7) | 18,211 (11.7) |  |
| Indian | 109,773 (20.7) | 109,773 (20.7) |  | 71,605 (18.7) | 71,605 (18.7) |  | 32,259 (20.7) | 32,259 (20.7) |  |
| Nepalese | 26,365 (5.0) | 26,365 (5.0) | 0.00 | 17,776 (4.6) | 17,776 (4.6) | 0.00 | 3,076 (2.0) | 3,076 (2.0) | 0.00 |
| Pakistani | 21,079 (4.0) | 21,079 (4.0) |  | 15,029 (3.9) | 15,029 (3.9) |  | 5,895 (3.8) | 5,895 (3.8) |  |
| Qatari | 125,512 (23.7) | 125,512 (23.7) |  | 98,159 (25.6) | 98,159 (25.6) |  | 39,767 (25.5) | 39,767 (25.5) |  |
| Sri Lankan | 12,600 (2.4) | 12,600 (2.4) |  | 8,913 (2.3) | 8,913 (2.3) |  | 2,549 (1.6) | 2,549 (1.6) |  |
| Sudanese | 15,096 (2.9) | 15,096 (2.9) |  | 11,181 (2.9) | 11,181 (2.9) |  | 3,965 (2.5) | 3,965 (2.5) |  |
| Other nationalities<sup>**</sup> | 104,643 (19.7) | 104,643 (19.7) |  | 82,117 (21.4) | 82,117 (21.4) |  | 36,780 (23.6) | 36,780 (23.6) |  |
| Number of coexisting conditions |  |  |  |  |  |  |  |  |  |
| None | 401,897 (75.8) | 401,897 (75.8) |  | 296,231 (77.4) | 296,231 (77.4) |  | 117,363 (75.2) | 117,363 (75.2) |  |
| 1 | 73,842 (13.9) | 73,842 (13.9) | 0.00 | 53,034 (13.9) | 53,034 (13.9) | 0.00 | 23,603 (15.1) | 23,603 (15.1) | 0.00 |
| 2 | 27,745 (5.2) | 27,745 (5.2) |  | 18,300 (4.8) | 18,300 (4.8) |  | 7,758 (5.0) | 7,758 (5.0) |  |
| ≥3 | 26,729 (5.0) | 26,729 (5.0) |  | 15,413 (4.0) | 15,413 (4.0) |  | 7,391 (4.7) | 7,391 (4.7) |  |

Abbreviations: IQR, interquartile range, SARS-CoV-2, severe acute respiratory syndrome coronavirus 2, and SMD standardized mean difference.
\*Cases and controls were matched exactly one-to-one by sex, 10-year age group, nationality, number of coexisting conditions, method of testing (PCR or rapid antigen), reason for SARS-CoV-2 testing, vaccine type, number of vaccine doses, and (by design) calendar month of testing.
<sup>†</sup>SMD is the difference in the mean of a covariate between groups divided by the pooled standard deviation. An SMD ≤0.1 indicates adequate matching.
<sup>‡</sup>Cases and controls were matched exactly one-to-one by sex, 10-year age group, nationality, number of coexisting conditions, method of testing (PCR or rapid antigen), reason for SARS-CoV-2 testing, status of most recent prior infection (no documented prior infection, documented pre-Omicron prior infection, or documented Omicron prior infection), and (by design) calendar month of testing.
<sup>§</sup>SMD is for the mean difference between groups divided by the pooled standard deviation.
<sup>¶</sup>Nationality groups were chosen to represent the most populous groups in Qatar.
<sup>\*\*</sup>Each case in the “other nationalities” group was matched, in a one-to-one ratio to a control with identical nationality. This group comprises up to 137 other nationalities in cases and controls.

**Table S3:**
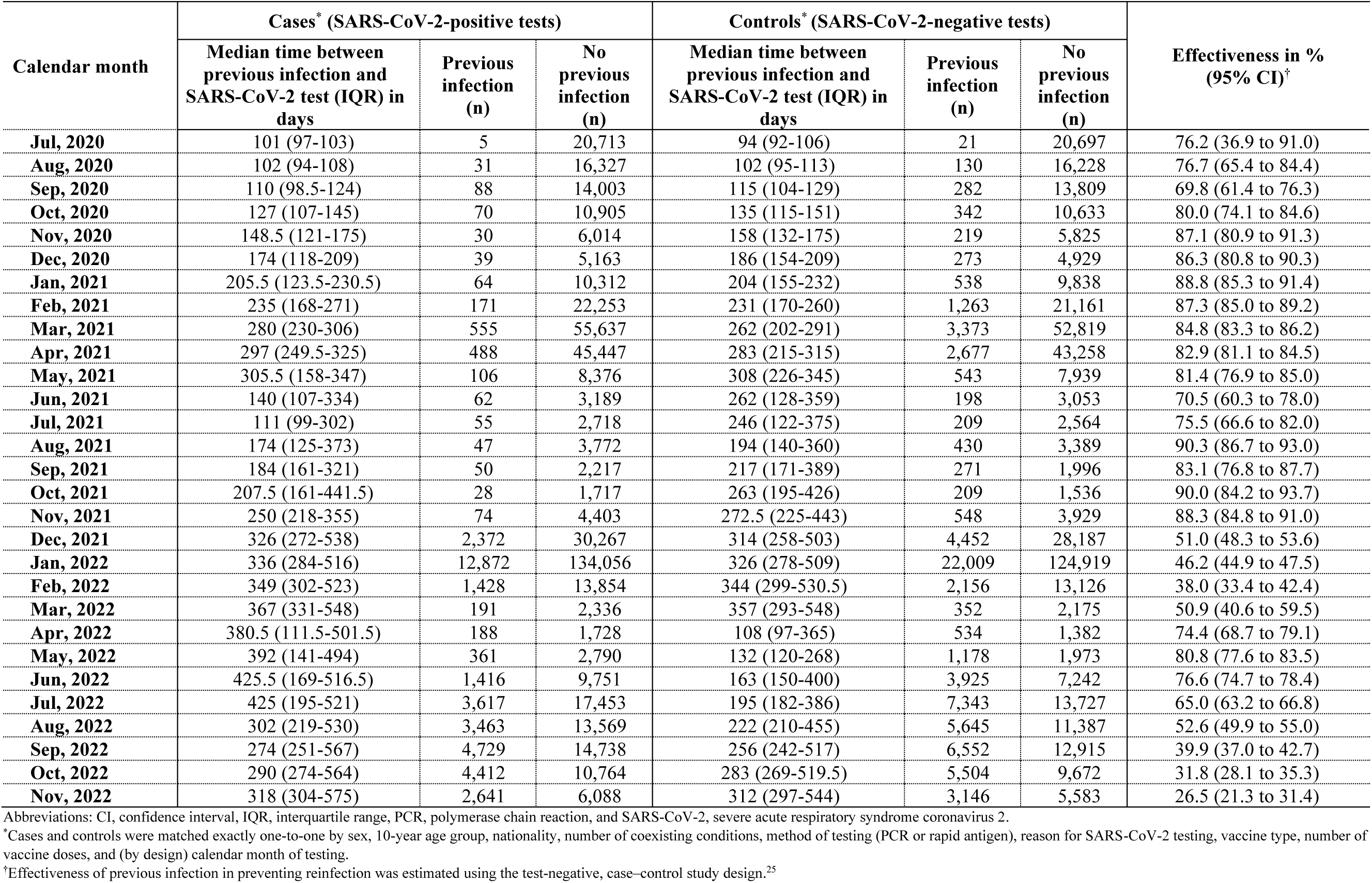
Effectiveness of previous SARS-CoV-2 infection against reinfection in Qatar between July of 2020 and November of 2022.

**Table S4:** Effectiveness of primary-series (two-dose) mRNA vaccination against SARS-CoV-2 infection in Qatar between February of 2021 and November of 2022.

| Calendar month | Cases* (SARS-CoV-2-positive tests) |  |  | Controls* (SARS-CoV-2-negative tests) |  |  | Effectiveness in %<br>(95% CI)<sup>†</sup> |
| --- | --- | --- | --- | --- | --- | --- | --- |
|  | Median time between<br>second vaccine dose and<br>SARS-CoV-2 test (IQR)<br>in days | Vaccinated<br>(n) | Not<br>vaccinated<br>(n) | Median time between<br>second vaccine dose and<br>SARS-CoV-2 test (IQR)<br>in days | Vaccinated<br>(n) | Not<br>vaccinated<br>(n) |  |
| Feb, 2021 | 24 (16-31) | 35 | 21,863 | 22 (17-28) | 221 | 21,677 | 84.5 (77.8 to 89.2) |
| Mar, 2021 | 34 (22-47) | 590 | 50,334 | 30 (20-43) | 2,394 | 48,530 | 81.8 (79.8 to 83.6) |
| Apr, 2021 | 34 (23-53) | 2,043 | 39,210 | 28 (20-44) | 7,569 | 33,684 | 84.0 (83.0 to 85.0) |
| May, 2021 | 48 (29-68.5) | 708 | 6,767 | 40 (26-59) | 2,016 | 5,459 | 80.9 (78.4 to 83.1) |
| Jun, 2021 | 64 (35-92) | 481 | 2,549 | 49 (31-79) | 1,151 | 1,879 | 80.7 (77.2 to 83.7) |
| Jul, 2021 | 107 (71-130) | 730 | 1,939 | 77 (51-114) | 1,258 | 1,411 | 76.2 (71.8 to 79.9) |
| Aug, 2021 | 134 (94-155) | 1,638 | 2,162 | 113 (74-150) | 2,158 | 1,642 | 63.0 (57.8 to 67.5) |
| Sep, 2021 | 157 (118-181) | 1,028 | 1,241 | 145 (97-176) | 1,316 | 953 | 61.9 (54.7 to 68.0) |
| Oct, 2021 | 180 (135-210) | 761 | 1,014 | 163 (114-204) | 889 | 886 | 44.6 (32.8 to 54.4) |
| Nov, 2021 | 209 (160-240) | 1,686 | 2,818 | 187 (133-230) | 2,096 | 2,408 | 52.7 (46.5 to 58.2) |
| Dec, 2021 | 228 (187-265) | 20,139 | 10,896 | 213 (163-251) | 19,963 | 11,072 | -3.9 (-7.8 to 0.0) |
| Jan, 2022 | 241 (192-279) | 77,910 | 51,637 | 237 (185-278) | 74,899 | 54,648 | -15.1 (-16.8 to -13.3) |
| Feb, 2022 | 248 (199-299) | 6,095 | 7,786 | 253 (198-302) | 6,052 | 7,829 | -2.2 (-8.2 to 4.0) |
| Mar, 2022 | 291 (236-347) | 1,025 | 1,060 | 282 (223-335) | 1,042 | 1,043 | 5.1 (-9.6 to 18.6) |
| Apr, 2022 | 326.5 (267-373) | 822 | 723 | 322 (261-371) | 789 | 756 | -12.4 (-26.5 to 4.2) |
| May, 2022 | 364 (289-412) | 1,206 | 1,092 | 344.5 (274-399.5) | 1,200 | 1,098 | -1.6 (-14.7 to 11.9) |
| Jun, 2022 | 389 (321-439) | 4,063 | 3,362 | 381 (313-434) | 3,773 | 3,652 | -20.7 (-26.7 to -14.2) |
| Jul, 2022 | 411 (346-460.5) | 7,528 | 7,237 | 409 (345-459) | 7,351 | 7,414 | -7.2 (-12.3 to -1.7) |
| Aug, 2022 | 432 (360-483) | 5,589 | 6,399 | 428 (356-483) | 5,568 | 6,420 | -1.1 (-7.1 to 5.1) |
| Sep, 2022 | 459 (393-510) | 6,990 | 6,186 | 460 (393-511) | 6,849 | 6,327 | -5.8 (-11.1 to -0.3) |
| Oct, 2022 | 500 (434-549) | 5,672 | 4,338 | 498 (433-548) | 5,550 | 4,460 | -6.9 (-12.9 to -0.4) |
| Nov, 2022 | 541 (486-585) | 3,064 | 2,562 | 533 (482-578) | 3,044 | 2,582 | -2.1 (-10.5 to 6.7) |

Abbreviations: CI, confidence interval, IQR, interquartile range, PCR, polymerase chain reaction, and SARS-CoV-2, severe acute respiratory syndrome coronavirus 2.
\*Cases and controls were matched exactly one-to-one by sex, 10-year age group, nationality, number of coexisting conditions, method of testing (PCR or rapid antigen), reason for SARS-CoV-2 testing, status of most recent prior infection (no documented prior infection, documented pre-Omicron prior infection, or documented Omicron prior infection), and (by design) calendar month of testing.
<sup>†</sup>Vaccine effectiveness was estimated using the test-negative, case-control study design.<sup>26,27</sup>

**Table S5:** Effectiveness of booster (third-dose) mRNA vaccination against SARS-CoV-2 infection in Qatar between November of 2021 and November of 2022.

| Calendar month | Cases* (SARS-CoV-2-positive tests) |  |  | Controls* (SARS-CoV-2-negative tests) |  |  | Effectiveness in %<br>(95% CI)<sup>†</sup> |
| --- | --- | --- | --- | --- | --- | --- | --- |
|  | Median time between<br>third vaccine dose and<br>SARS-CoV-2 test (IQR)<br>in days | Vaccinated<br>(n) | Not<br>vaccinated<br>(n) | Median time between<br>third vaccine dose and<br>SARS-CoV-2 test (IQR)<br>in days | Vaccinated<br>(n) | Not<br>vaccinated<br>(n) |  |
| **Nov, 2021** | 27 (8.5-47) | 16 | 2,770 | 20 (12-34) | 60 | 2,726 | 83.0 (65.6 to 91.6) |
| **Dec, 2021** | 34 (20-52) | 1,948 | 10,541 | 26 (16-41) | 2,354 | 10,135 | 32.9 (26.7 to 38.5) |
| **Jan, 2022** | 43 (26-60) | 15,607 | 52,535 | 41 (24-57) | 16,996 | 51,146 | 18.1 (15.4 to 20.8) |
| **Feb, 2022** | 63 (38-82) | 1,591 | 7,719 | 60 (37-81) | 1,631 | 7,679 | 5.3 (-4.7 to 14.6) |
| **Mar, 2022** | 97.5 (64-116) | 484 | 1,051 | 80 (48-111) | 495 | 1,040 | 6.0 (-13.6 to 23.7) |
| **Apr, 2022** | 123 (85-144) | 422 | 715 | 101 (69-136) | 397 | 740 | -15.6 (-32.9 to 5.7) |
| **May, 2022** | 158 (125-178) | 864 | 1,090 | 135 (96-167) | 823 | 1,131 | -13.7 (-27.0 to 1.9) |
| **Jun, 2022** | 181 (148-204) | 3,827 | 3,343 | 164 (125-195) | 3,238 | 3,932 | -42.8 (-47.6 to -37.6) |
| **Jul, 2022** | 204 (170-231) | 6,110 | 7,214 | 193 (151-226) | 5,665 | 7,659 | -21.5 (-26.4 to -16.2) |
| **Aug, 2022** | 228 (190-257) | 4,952 | 6,361 | 215 (169-251) | 4,682 | 6,631 | -17.4 (-23.3 to -11.1) |
| **Sep, 2022** | 257 (217-288) | 6,017 | 6,140 | 245 (201-283) | 5,749 | 6,408 | -15.1 (-20.7 to -9.1) |
| **Oct, 2022** | 286 (245-316) | 4,979 | 4,305 | 278 (233-313) | 4,692 | 4,592 | -19.8 (-25.8 to -13.4) |
| **Nov, 2022** | 317 (278-349) | 2,973 | 2,541 | 307 (264-342) | 2,715 | 2,799 | -28.0 (-34.9 to -20.5) |

Abbreviations: CI, confidence interval, IQR, interquartile range, PCR, polymerase chain reaction, and SARS-CoV-2, severe acute respiratory syndrome coronavirus 2.
\*Cases and controls were matched exactly one-to-one by sex, 10-year age group, nationality, number of coexisting conditions, method of testing (PCR or rapid antigen), reason for SARS-CoV-2 testing, status of most recent prior infection (no documented prior infection, documented pre-Omicron prior infection, or documented Omicron prior infection), and (by design) calendar month of testing.
<sup>†</sup>Vaccine effectiveness was estimated using the test-negative, case-control study design.<sup>26,27</sup>

**Table S6:** Effectiveness against severe, critical, or fatal COVID-19 of A) previous SARS-CoV-2 infection, B) primary-series (two-dose) mRNA vaccination, and C) booster (third-dose) mRNA vaccination, in Qatar, between July of 2020 and November of 2022.

| A) Effectiveness of previous SARS-CoV-2 infection |  |  |  |  |  |  |  |
| --- | --- | --- | --- | --- | --- | --- | --- |
| Calendar month | Cases* (SARS-CoV-2-positive tests) |  |  | Controls* (SARS-CoV-2-negative tests) |  |  | Effectiveness in % (95% CI)<sup>‡</sup> |
|  | Median time between previous infection and SARS-CoV-2 test (IQR) in days | Previous infection (n) | No previous infection (n) | Median time between previous infection and SARS-CoV-2 test (IQR) in days | Previous infection (n) | No previous infection (n) |  |
| Jul-Dec, 2020 | 196 (196-196) | 1 | 1,582 | 138 (110-172) | 195 | 7,460 | 97.6 (82.8 to 99.7) |
| Jan-Jun, 2021 | 280 (227-296) | 10 | 5,354 | 244 (191-288) | 1,777 | 23,479 | 97.6 (95.5 to 98.7) |
| Jul-Dec, 2021 | 554.5 (514-595) | 2 | 360 | 237 (60-376) | 225 | 1,347 | 96.8 (87.0 to 99.2) |
| Jan-Jun, 2022 | 294 (263.5-395.5) | 4 | 280 | 350 (277.5-508.5) | 232 | 967 | 94.1 (83.9 to 97.8) |
| Jul-Nov, 2022 | 610 (550-670) | 2 | 29 | 217 (182.5-319) | 60 | 78 | 91.1 (60.5 to 98.0) |
| B) Effectiveness of primary-series (two-dose) mRNA vaccination |  |  |  |  |  |  |  |
| Calendar month | Cases<sup>†</sup> (SARS-CoV-2-positive tests) |  |  | Controls<sup>†</sup> (SARS-CoV-2-negative tests) |  |  | Effectiveness in % (95% CI)<sup>‡</sup> |
|  | Median time between second vaccine dose and SARS-CoV-2 test (IQR) in days | Vaccinated (n) | Not vaccinated (n) | Median time between second vaccine dose and SARS-CoV-2 test (IQR) in days | Vaccinated (n) | Not vaccinated (n) |  |
| Feb-Apr, 2021 | 49.5 (37-71) | 42 | 4,085 | 31 (21-47) | 2,749 | 16,974 | 95.4 (93.6 to 96.7) |
| May-Jul, 2021 | 95 (79-107) | 25 | 386 | 64 (38-95) | 1,087 | 833 | 96.9 (94.8 to 98.1) |
| Aug-Oct, 2021 | 189 (157.5-210.5) | 60 | 115 | 159 (116-190) | 649 | 156 | 91.4 (86.6 to 94.5) |
| Nov, 2021-Jan, 2022 | 261 (202-304) | 161 | 206 | 242 (193-285) | 1,217 | 394 | 82.3 (76.6 to 86.6) |
| Feb-Apr, 2022 | 254 (225-308) | 9 | 18 | 313.5 (275-356.5) | 76 | 35 | 81.4 (48.5 to 93.3) |
| May-Jul, 2022 | 425 (314-447) | 3 | 8 | 431 (371-465) | 33 | 19 | 83.8 (14.1 to 96.9) |
| Aug-Nov, 2022 | 453 (408-566) | 7 | 10 | 511 (442-564) | 39 | 32 | 53.1 (-41.3 to 87.1) |
| C) Effectiveness of booster (third-dose) mRNA vaccination |  |  |  |  |  |  |  |
| Calendar month | Cases<sup>†</sup> (SARS-CoV-2-positive tests) |  |  | Controls<sup>†</sup> (SARS-CoV-2-negative tests) |  |  | Effectiveness in % (95% CI)<sup>‡</sup> |
|  | Median time between third vaccine dose and SARS-CoV-2 test (IQR) in days | Vaccinated (n) | Not vaccinated (n) | Median time between third vaccine dose and SARS-CoV-2 test (IQR) in days | Vaccinated (n) | Not vaccinated (n) |  |
| Nov, 2021-Jan, 2022 | 84 (49-113) | 22 | 203 | 44 (23-75) | 460 | 508 | 95.0 (90.5 to 97.4) |
| Feb-Apr, 2022 | 18.5 (9-28) | 2 | 17 | 96 (72-127) | 45 | 33 | 96.2 (69.6 to 99.5) |
| **May-Jul, 2022** | 225 (183-269) | 6 | 8 | 195 (139-218) | 33 | 27 | 43.6 (-51.4 to 84.5) |
| **Aug-Nov, 2022** | 328.5 (255.5-343) | 4 | 10 | 276 (219-306) | 30 | 24 | 90.4 (9.6 to 99.0) |

Abbreviations: CI, confidence interval, COVID-19, coronavirus disease 2019, IQR, interquartile range, PCR, polymerase chain reaction, and SARS-CoV-2, severe acute respiratory syndrome coronavirus 2.
\*Cases and controls were matched exactly one-to-one by sex, 10-year age group, nationality, number of coexisting conditions, method of testing (PCR or rapid antigen), reason for SARS-CoV-2 testing, vaccine type, number of vaccine doses, and (by design) 6 calendar months of testing.
†Cases and controls were matched exactly one-to-one by sex, 10-year age group, nationality, number of coexisting conditions, method of testing (PCR or rapid antigen), reason for SARS-CoV-2 testing, status of most recent prior infection (no documented prior infection, documented pre-Omicron prior infection, or documented Omicron prior infection), and (by design) 6 calendar months of testing.
‡Effectiveness of previous infection in preventing reinfection and vaccine effectiveness were estimated using the test-negative, case-control study design.<sup>25-27</sup>

## References

1. Abu-Raddad LJ, Chemaitelly H, Butt AA, National Study Group for Covid Vaccination. Effectiveness of the BNT162b2 Covid-19 Vaccine against the B.1.1.7 and B.1.351 Variants. N Engl J Med 2021; 385(2): 187–9.

2. Chemaitelly H, Yassine HM, Benslimane FM, et al. mRNA-1273 COVID-19 vaccine effectiveness against the B.1.1.7 and B.1.351 variants and severe COVID-19 disease in Qatar. Nat Med 2021; 27(9): 1614–21.

3. Chemaitelly H, Tang P, Hasan MR, et al. Waning of BNT162b2 Vaccine Protection against SARS-CoV-2 Infection in Qatar. N Engl J Med 2021; 385(24): e83.

4. Abu-Raddad LJ, Chemaitelly H, Bertollini R, National Study Group for Covid Vaccination. Waning mRNA-1273 Vaccine Effectiveness against SARS-CoV-2 Infection in Qatar. N Engl J Med 2022; 386(11): 1091–3.

5. Feikin DR, Higdon MM, Abu-Raddad LJ, et al. Duration of effectiveness of vaccines against SARS-CoV-2 infection and COVID-19 disease: results of a systematic review and meta-regression. Lancet 2022.

6. Higdon MM, Baidya A, Walter KK, et al. Duration of effectiveness of vaccination against COVID-19 caused by the omicron variant. Lancet Infect Dis 2022; 22(8): 1114–6.

7. Andrews N, Stowe J, Kirsebom F, et al. Covid-19 Vaccine Effectiveness against the Omicron (B.1.1.529) Variant. N Engl J Med 2022; 386(16): 1532–46.

8. Chemaitelly H, Ayoub HH, AlMukdad S, et al. Duration of mRNA vaccine protection against SARS-CoV-2 Omicron BA.1 and BA.2 subvariants in Qatar. Nat Commun 2022; 13(1): 3082.

9. Abu-Raddad LJ, Chemaitelly H, Ayoub HH, et al. Effect of mRNA Vaccine Boosters against SARS-CoV-2 Omicron Infection in Qatar. N Engl J Med 2022; 386(19): 1804–16.

10. Chemaitelly H, Ayoub HH, Tang P, et al. Long-term COVID-19 booster effectiveness by infection history and clinical vulnerability and immune imprinting: a retrospective population-based cohort study. The Lancet Infectious Diseases 2023.

11. Abu-Raddad LJ, Chemaitelly H, Malek JA, et al. Assessment of the Risk of Severe Acute Respiratory Syndrome Coronavirus 2 (SARS-CoV-2) Reinfection in an Intense Reexposure Setting. Clin Infect Dis 2021; 73(7): e1830–e40.

12. Abu-Raddad LJ, Chemaitelly H, Coyle P, et al. SARS-CoV-2 antibody-positivity protects against reinfection for at least seven months with 95% efficacy. EClinicalMedicine 2021; 35: 100861.

13. Chemaitelly H, Nagelkerke N, Ayoub HH, et al. Duration of immune protection of SARS-CoV-2 natural infection against reinfection. J Travel Med 2022; 29(8).

14. Altarawneh HN, Chemaitelly H, Ayoub HH, et al. Effects of Previous Infection and Vaccination on Symptomatic Omicron Infections. N Engl J Med 2022; 387(1): 21–34.

15. Altarawneh HN, Chemaitelly H, Hasan MR, et al. Protection against the Omicron Variant from Previous SARS-CoV-2 Infection. N Engl J Med 2022; 386(13): 1288–90.

16. Chemaitelly H, Tang P, Coyle P, et al. Protection against Reinfection with the Omicron BA.2.75 Subvariant. N Engl J Med 2023; 388(7): 665–7.

17. Chemaitelly H, Ayoub HH, Coyle P, et al. Protection of Omicron sub-lineage infection against reinfection with another Omicron sub-lineage. Nat Commun 2022; 13(1): 4675.

18. Altarawneh HN, Chemaitelly H, Ayoub HH, et al. Protective Effect of Previous SARS-CoV-2 Infection against Omicron BA.4 and BA.5 Subvariants. N Engl J Med 2022; 387(17): 1620–2.

19. Abu-Raddad LJ, Chemaitelly H, Ayoub HH, et al. Characterizing the Qatar advanced-phase SARS-CoV-2 epidemic. Sci Rep 2021; 11(1): 6233.

20. Abu-Raddad LJ, Chemaitelly H, Ayoub HH, et al. Introduction and expansion of the SARS-CoV-2 B.1.1.7 variant and reinfections in Qatar: A nationally representative cohort study. PLoS Med 2021; 18(12): e1003879.

21. Chemaitelly H, Bertollini R, Abu-Raddad LJ, National Study Group for Covid Epidemiology. Efficacy of Natural Immunity against SARS-CoV-2 Reinfection with the Beta Variant. N Engl J Med 2021; 385(27): 2585–6.

22. Tang P, Hasan MR, Chemaitelly H, et al. BNT162b2 and mRNA-1273 COVID-19 vaccine effectiveness against the SARS-CoV-2 Delta variant in Qatar. Nat Med 2021; 27(12): 2136–43.

23. Abu-Raddad LJ, Chemaitelly H, Bertollini R, National Study Group for Covid Vaccination. Effectiveness of mRNA-1273 and BNT162b2 Vaccines in Qatar. N Engl J Med 2022; 386(8): 799–800.

24. Chemaitelly H, Faust JS, Krumholz H, et al. Short- and longer-term all-cause mortality among SARS-CoV-2-infected persons and the pull-forward phenomenon in Qatar. medRxiv 2023: 2023.01.29.23285152.

25. Ayoub HH, Tomy M, Chemaitelly H, et al. Estimating protection afforded by prior infection in preventing reinfection: Applying the test-negative study design. medRxiv 2022: 2022.01.02.22268622.

26. Jackson ML, Nelson JC. The test-negative design for estimating influenza vaccine effectiveness. Vaccine 2013; 31(17): 2165–8.

27. Verani JR, Baqui AH, Broome CV, et al. Case-control vaccine effectiveness studies: Preparation, design, and enrollment of cases and controls. Vaccine 2017; 35(25): 3295–302.

28. Lopez Bernal J, Andrews N, Gower C, et al. Effectiveness of Covid-19 Vaccines against the B.1.617.2 (Delta) Variant. N Engl J Med 2021; 385(7): 585–94.

29. Ayoub HH, Chemaitelly H, Seedat S, et al. Mathematical modeling of the SARS-CoV-2 epidemic in Qatar and its impact on the national response to COVID-19. J Glob Health 2021; 11: 05005.

30. Coyle PV, Chemaitelly H, Ben Hadj Kacem MA, et al. SARS-CoV-2 seroprevalence in the urban population of Qatar: An analysis of antibody testing on a sample of 112,941 individuals. iScience 2021; 24(6): 102646.

31. Al-Thani MH, Farag E, Bertollini R, et al. SARS-CoV-2 Infection Is at Herd Immunity in the Majority Segment of the Population of Qatar. Open Forum Infect Dis 2021; 8(8): ofab221.

32. Jeremijenko A, Chemaitelly H, Ayoub HH, et al. Herd Immunity against Severe Acute Respiratory Syndrome Coronavirus 2 Infection in 10 Communities, Qatar. Emerg Infect Dis 2021; 27(5): 1343–52.

33. Abu-Raddad LJ, Chemaitelly H, Yassine HM, et al. Pfizer-BioNTech mRNA BNT162b2 Covid-19 vaccine protection against variants of concern after one versus two doses. J Travel Med 2021; 28(7).

34. Pilz S, Theiler-Schwetz V, Trummer C, Krause R, Ioannidis JPA. SARS-CoV-2 reinfections: Overview of efficacy and duration of natural and hybrid immunity. Environ Res 2022: 112911.

35. Kojima N, Shrestha NK, Klausner JD. A Systematic Review of the Protective Effect of Prior SARS-CoV-2 Infection on Repeat Infection. Eval Health Prof 2021; 44(4): 327–32.

36. World Health Organization (WHO). Living guidance for clinical management of COVID-19. Aavailable from: https://www.who.int/publications/i/item/WHO-2019-nCoV-clinical-2021-2. Accessed on: February 27, 2023. 2021.

37. World Health Organization (WHO). International Guidelines for Certification and Classification (Coding) of COVID-19 as Cause of Death. Available from: https://www.who.int/publications/m/item/international-guidelines-for-certification-and-classification-(coding)-of-covid-19-as-cause-of-death. Accessed on: February 27, 2023. 2020.

38. Austin PC. Using the Standardized Difference to Compare the Prevalence of a Binary Variable Between Two Groups in Observational Research. Communications in Statistics - Simulation and Computation 2009; 38(6): 1228–34.

39. Jacoby P, Kelly H. Is it necessary to adjust for calendar time in a test negative design?: Responding to: Jackson ML, Nelson JC. The test negative design for estimating influenza vaccine effectiveness. Vaccine 2013;31(April (17)):2165–8. Vaccine 2014; 32(25): 2942.

40. Pearce N. Analysis of matched case-control studies. BMJ 2016; 352: i969.

41. Rothman KJ, Greenland S, Lash TL. Modern epidemiology. 3rd ed. Philadelphia: Wolters Kluwer Health/Lippincott Williams & Wilkins; 2008.

42. Tseng HF, Ackerson BK, Bruxvoort KJ, et al. Effectiveness of mRNA-1273 vaccination against SARS-CoV-2 omicron subvariants BA.1, BA.2, BA.2.12.1, BA.4, and BA.5. Nat Commun 2023; 14(1): 189.

43. Subissi L, von Gottberg A, Thukral L, et al. An early warning system for emerging SARS-CoV-2 variants. Nat Med 2022; 28(6): 1110–5.

44. Kissler SM, Tedijanto C, Goldstein E, Grad YH, Lipsitch M. Projecting the transmission dynamics of SARS-CoV-2 through the postpandemic period. Science 2020; 368(6493): 860–8.

45. Patel MM, York IA, Monto AS, Thompson MG, Fry AM. Immune-mediated attenuation of influenza illness after infection: opportunities and challenges. Lancet Microbe 2021; 2(12): e715–e25.

46. Lavine JS, Bjornstad ON, Antia R. Immunological characteristics govern the transition of COVID-19 to endemicity. Science 2021; 371(6530): 741–5.

47. Chemaitelly H, Ayoub HH, Tang P, et al. COVID-19 primary series and booster vaccination and immune imprinting. medRxiv 2022: 2022.10.31.22281756.

48. Janjua NZ, Skowronski DM, Hottes TS, et al. Seasonal influenza vaccine and increased risk of pandemic A/H1N1-related illness: first detection of the association in British Columbia, Canada. Clin Infect Dis 2010; 51(9): 1017–27.

49. Skowronski DM, Sabaiduc S, Leir S, et al. Paradoxical clade- and age-specific vaccine effectiveness during the 2018/19 influenza A(H3N2) epidemic in Canada: potential imprint-regulated effect of vaccine (I-REV). Euro Surveill 2019; 24(46).

50. Ray GT, Lewis N, Klein NP, Daley MF, Lipsitch M, Fireman B. Depletion-of-susceptibles Bias in Analyses of Intra-season Waning of Influenza Vaccine Effectiveness. Clin Infect Dis 2020; 70(7): 1484–6.

## References

1. Altarawneh HN, Chemaitelly H, Ayoub HH, et al. Effects of Previous Infection and Vaccination on Symptomatic Omicron Infections. N Engl J Med 2022; 387(1): 21–34.

2. Chemaitelly H, Tang P, Hasan MR, et al. Waning of BNT162b2 Vaccine Protection against SARS-CoV-2 Infection in Qatar. N Engl J Med 2021; 385(24): e83.

3. Altarawneh HN, Chemaitelly H, Hasan MR, et al. Protection against the Omicron Variant from Previous SARS-CoV-2 Infection. N Engl J Med 2022; 386(13): 1288–90.

4. Planning and Statistics Authority-State of Qatar. Qatar Monthly Statistics. Available from: https://www.psa.gov.qa/en/pages/default.aspx. Accessed on: May 26, 2020. 2020.

5. Abu-Raddad LJ, Chemaitelly H, Ayoub HH, et al. Characterizing the Qatar advanced-phase SARS-CoV-2 epidemic. Sci Rep 2021; 11(1): 6233.

6. Chemaitelly H, Bertollini R, Abu-Raddad LJ, National Study Group for Covid Epidemiology. Efficacy of Natural Immunity against SARS-CoV-2 Reinfection with the Beta Variant. N Engl J Med 2021; 385(27): 2585–6.

7. Abu-Raddad LJ, Chemaitelly H, Ayoub HH, et al. Effect of mRNA Vaccine Boosters against SARS-CoV-2 Omicron Infection in Qatar. N Engl J Med 2022; 386(19): 1804–16.

8. Vogels C, Fauver J, Grubaugh N. Multiplexed RT-qPCR to screen for SARS-COV-2 B.1.1.7, B.1.351, and P.1 variants of concern V.3. dx.doi.org/10.17504/protocols.io.br9vm966. 2021; (June 6, 2021).

9. Abu-Raddad LJ, Chemaitelly H, Butt AA, National Study Group for Covid Vaccination. Effectiveness of the BNT162b2 Covid-19 Vaccine against the B.1.1.7 and B.1.351 Variants. N Engl J Med 2021; 385(2): 187–9.

10. Chemaitelly H, Yassine HM, Benslimane FM, et al. mRNA-1273 COVID-19 vaccine effectiveness against the B.1.1.7 and B.1.351 variants and severe COVID-19 disease in Qatar. Nat Med 2021; 27(9): 1614–21.

11. National Project of Surveillance for Variants of Concern and Viral Genome Sequencing. Qatar viral genome sequencing data. Data on randomly collected samples. https://www.gisaid.org/phylodynamics/global/nextstrain/. 2021. https://www.gisaid.org/phylodynamics/global/nextstrain/.

12. Benslimane FM, Al Khatib HA, Al-Jamal O, et al. One Year of SARS-CoV-2: Genomic Characterization of COVID-19 Outbreak in Qatar. Front Cell Infect Microbiol 2021; 11: 768883.

13. Hasan MR, Kalikiri MKR, Mirza F, et al. Real-Time SARS-CoV-2 Genotyping by High-Throughput Multiplex PCR Reveals the Epidemiology of the Variants of Concern in Qatar. Int J Infect Dis 2021; 112: 52–4.

14. Saththasivam J, El-Malah SS, Gomez TA, et al. COVID-19 (SARS-CoV-2) outbreak monitoring using wastewater-based epidemiology in Qatar. Sci Total Environ 2021; 774: 145608.

15. El-Malah SS, Saththasivam J, Jabbar KA, et al. Application of human RNase P normalization for the realistic estimation of SARS-CoV-2 viral load in wastewater: A perspective from Qatar wastewater surveillance. Environ Technol Innov 2022; 27: 102775.

16. Tang P, Hasan MR, Chemaitelly H, et al. BNT162b2 and mRNA-1273 COVID-19 vaccine effectiveness against the SARS-CoV-2 Delta variant in Qatar. Nat Med 2021; 27(12): 2136–43.

17. Chemaitelly H, Ayoub HH, AlMukdad S, et al. Duration of mRNA vaccine protection against SARS-CoV-2 Omicron BA.1 and BA.2 subvariants in Qatar. Nat Commun 2022; 13(1): 3082.

18. Qassim SH, Chemaitelly H, Ayoub HH, et al. Effects of BA.1/BA.2 subvariant, vaccination and prior infection on infectiousness of SARS-CoV-2 omicron infections. J Travel Med 2022; 29(6).

19. Altarawneh HN, Chemaitelly H, Ayoub HH, et al. Protective Effect of Previous SARS-CoV-2 Infection against Omicron BA.4 and BA.5 Subvariants. N Engl J Med 2022; 387(17): 1620–2.

20. Chemaitelly H, Tang P, Coyle P, et al. Protection against Reinfection with the Omicron BA.2.75 Subvariant. N Engl J Med 2023; 388(7): 665–7.

21. World Health Organization (WHO). Living guidance for clinical management of COVID-19. Aavailable from: https://www.who.int/publications/i/item/WHO-2019-nCoV-clinical-2021-2. Accessed on: February 27, 2023. 2021.

22. World Health Organization (WHO). International Guidelines for Certification and Classification (Coding) of COVID-19 as Cause of Death. Available from: https://www.who.int/publications/m/item/international-guidelines-for-certification-and-classification-(coding)-of-covid-19-as-cause-of-death. Accessed on: February 27, 2023. 2020.

